# Health-related quality of life among patients with sepsis in Blantyre, Malawi: an observational cohort study

**DOI:** 10.1101/2025.06.04.25328855

**Authors:** Nateiya M Yongolo, Joseph M Lewis, Laura Rosu, Ibrahim Simiyu, Grace Katha, Jane Mallewa, Stephen A Spencer, Gift T Banda, Sangwani N. Salimu, Ben Morton, Rachel Manongi, Jamie Rylance, Nicholas A Feasey, Matthew P. Rubach, Eve Worrall

## Abstract

Sepsis is a life-threatening condition with high mortality, especially in sub-Saharan Africa (SSA). Sepsis survivors may face long term consequences and experience poor health-related quality of life (HRQoL). By comparing HRQoL trajectories between sepsis cases and matched controls we sought to identify the longitudinal impact of sepsis on HRQoL in the SSA context.

This study was nested within a longitudinal adult sepsis cohort admitted to hospital in Blantyre, Malawi. Two reference control groups matched to cases by age, sex, and geographical location were recruited: (1) hospital inpatients without sepsis and (2) community controls with no current illness. All participants were followed up to 180 days post-enrolment. HRQoL was assessed using EQ-5D-3L questionnaire and a visual analogue scale (VAS). Regression analysis was conducted to examine factors associated with HRQoL among sepsis survivors.

A total of 425 participants: 225 sepsis cases, 100 hospital controls, and 100 community controls were recruited. HIV prevalence was higher among sepsis cases 143/225 (63.6%) compared to hospital controls 12/100 (12.0%) and community controls 18/100 (18.0%) p= <0.001. At baseline, sepsis cases had lower health utility scores (median 0.596, IQR: 0.365– 0.734 compared to hospital controls (0.666, IQR: 0.611 - 0.722) and community controls (0.900, IQR: 0.833 - 0.900). Over time, sepsis cases displayed fluctuating HRQoL, with a marked decline in utility scores at day 180 (0 IQR: 0 - 0) compared to relatively stable scores in both control groups. Regression analysis identified age, sex, duration of illness before admission and baseline utility score as significant predictors of HRQoL in the sepsis group. The findings reveal a severe and persistent reduction in HRQoL among patients admitted with sepsis in Malawi, suggesting a substantial post-discharge burden among survivors. These results point to a need for evidence-based prevention, early recognition of sepsis and post-sepsis support programs in SSA.

## Introduction

Sepsis is a life-threatening condition causing over 11 million deaths annually, with the highest burden in low and middle-income countries (LMICs), particularly in sub-Saharan Africa (SSA), yet its long-term consequences remain poorly understood in these settings (1–3). Sepsis leads to significant short and long-term morbidity among survivors. Beyond the acute phase, individuals often experience persistent physical, psychological, and functional impairments that compromise their health-related quality of life (HRQoL) ((1,2). In low-resource settings like Malawi, where sepsis prevalence among hospitalized patients ranges from 7.1% to 24% (4,5), and where young adults living with HIV are commonly affected (6,7), these consequences may be especially pronounced.

While global evidence increasingly recognizes long-term impairments after sepsis, most data come from high-income settings (8–11). In sub-Saharan Africa (SSA), there is limited understanding of post-sepsis outcomes, particularly regarding HRQoL. In Malawi late post-hospital mortality among sepsis patients has been reported, suggesting ongoing functional decline and compromised quality of life (6,7). However, empirical data to quantify this burden and explore contributing factors are lacking.

Several HRQoL assessment tools have been developed, many of which have been adapted into multiple languages (12). The EQ-5D-3L is particularly well-suited for economic evaluations, as it enables the calculation of quality-adjusted life years (QALYs), a metric that allow self-reporting and integrates both HRQoL (quality) and survival duration (quantity). The tool is a generic measure and not diseases specific and is suitable for sepsis which is syndromic condition with multiple underlying aetiologies.

This study estimates the impact of sepsis on HRQoL over six months using EQ-5D-3L, a methodology consistent with other studies (13,14). We compare HRQoL trajectories among sepsis cases, hospitalized controls without sepsis, and community controls, while also identifying key determinants of long-term HRQoL outcomes among sepsis cases. By addressing a critical knowledge gap in SSA, particularly in Malawi, our findings will inform post-sepsis rehabilitation strategies, with a focus on improving HRQoL outcomes.

In this study, we use data from a prospective cohort of adult patients admitted with sepsis at Queen Elizabeth Central Hospital in Blantyre, Malawi, to assess HRQoL over six months following hospitalisation. We compare outcomes between sepsis survivors, matched non-sepsis hospital controls, and matched community controls. Our primary aim is to describe HRQoL trajectories and identify factors associated with poor HRQoL in sepsis survivors. We hypothesize that sepsis cases have significantly lower HRQoL compared to hospitalized non-sepsis patients and community controls, with the greatest impairment observed among sepsis survivors over time. The general community reflects better HRQoL outcomes, serving as a useful comparator for sepsis-related disability.

## Materials and methods

### Study setting and design

This study was conducted at Queen Elizabeth Central Hospital (QECH) in Blantyre, Malawi, a government-operated tertiary referral hospital with approximately 1,350 beds. The hospital serves as the primary teaching hospital and provides both inpatient and outpatient services for Blantyre’s population The study was a prospective observational cohort study conducted from February 2017 to January 2019.This study is a Health Economic (HE) analysis conducted as a follow-up and using data from a larger observational cohort study (parent study) on sepsis in Malawi (6). The parent study aimed to characterize sepsis in Malawi, including causative pathogens, treatment administered, and 180-day mortality outcomes.

Prior publications have detailed the aetiology, clinical characteristics, and laboratory findings of sepsis patients in this cohort study (15). The study was conducted in accordance with recommended guidelines for conducting and reporting HRQoL by self-reported measures(16) and check list is provided as (S1 checklist).

### Study population

The cohort study was carried out among patients admitted through the emergency department of the QECH. The study included three groups: sepsis cases (group 1: sepsis cases), non-sepsis inpatients (group 2: hospital controls) and community members (group 3: community controls). The inclusion and exclusion criteria are presented (Table 1). The control groups were also matched 1:1 by age (±5 years), sex, to group 1 (sepsis cases), and for community controls also geographic location was matched.

**Table 1:** Eligibility criteria for study participants.

| **Group** | **Inclusion Criteria** | **Exclusion Criteria** |
| --- | --- | --- |
| **Sepsis Cases (Group 1)** | - Patients $\geq 16$ years presenting at QECH Emergency Department<br>- Evidence of infection (fever $>37.5^{\circ}\text{C}$ or history of fever in past 72 hours)<br>- At least one organ dysfunction marker<br>- Oxygen saturation $<90\%$<br>- Respiratory rate $>30$ breaths/min<br>- Systolic BP $<90$ mmHg<br>- Glasgow Coma Scale (GCS) $<15$ | - Unable to provide informed consent (no available guardian)<br>- Unable to speak Chichewa or English<br>- Residing $>30$ km from Blantyre<br>- Clinical decision to discontinue active care |
| **Non-Sepsis Inpatients (Hospital Controls, Group 2)** | - Patients $\geq 16$ years admitted to QECH medical wards<br>- No sepsis diagnosis (did not meet sepsis criteria)<br>- No antimicrobial therapy exposure before admission | - Same as Group 1 |
| **Community Controls (Group 3)** | - Individuals $\geq 16$ years from the same geographic area as Group 1<br>- No suspicion of infection in the past four weeks<br>- No recent antimicrobial therapy (except cotrimoxazole preventive therapy CPT or anti-tuberculosis (TB) chemotherapy) | - Unable to provide informed consent<br>- Unable to speak Chichewa or English |

### Study recruitment procedures

Recruitment occurred at QECH’s emergency department and medical Wards from Monday to Friday, 07:00–17:00. Patients were screened in emergency department as early in attendance as possible, where consecutive recruitment of sepsis cases (group 1) at the emergency was done daily while matching list was generated for Non sepsis inpatient controls (group 2) where matching participants were listed from the emergence attendees and then recruited after consenting and fitting the criteria. After explaining the aim of the study and associated study procedures, research assistants sought written consent from participants or legal guardians for proxy consent. Sepsis cases (group 1) were screened and recruited at the Emergency Department. Hospital controls (group 2) were also recruited from the Emergency Department. Community controls (group 3) were identified by random geographic matching, starting from a sepsis participant’s household and proceeding in a randomly selected direction (determined by spinning a bottle).

Sepsis cases (group 1) and hospital Controls (group 2) were assessed at: baseline (day 0), day 7, day 28, day 90 and day 180. Community Controls (group 3) were assessed at: baseline (day 0), day 28 and day 180. Making a total of 5 assessments for Sepsis cases, hospital controls and 3 assessments for community controls. A variation of plus and minus 3 days was considered HRQoL was assessed using the EQ-5D-3L questionnaire (17), which has been validated in Chichewa and approved by the EuroQol Research Foundation, a non-profit organisation based in the Netherlands dedicated to the development and application of standardized measures for HRQoL. The tool consists of a five dimensions health status index: mobility, self-care, usual activities, pain/discomfort and anxiety/depression. Each dimension was rated on a three-level severity scale: Level 1: No problem; Level 2: Some problems; and Level 3: Severe problems. The tool is accompanied by a Visual Analogue Scale (VAS) which is a self-rated health score from 0 (worst health) to 100 (best health). In LMICs, generic HRQoL tools such as the EQ-5D-3L and Short Form 36 (SF-36) (18–22) are recommended due to their ease of administration and cross-population comparability (23). These instruments have been widely used in health economic (HE) evaluations, facilitating comparisons of health gains and losses.

### Data collection and handling

Clinical, demographic, and socioeconomic data were collected using standardized electronic data collection forms using Open Data Kit (ODK)(24). Data were collected via face-to-face interviews; the research assistant administered the EQ-5D-3L tool using hardcopy questionnaires. Research assistants would ask the questions and allow participants to respond, and individual scores for all dimensions of the EQ-5D-3L were recorded. Participants were asked to indicate their current health status by marking a point on the EQ-5D Visual Analogue Scale (VAS), a vertical scale ranging from 0 (’the worst health you can imagine’) to 100 (’the best health you can imagine’). Investigations to determine the aetiology of sepsis were performed as described in (15); HIV infection status and prior or current tuberculosis were ascertained at enrolment.

The data were captured on the Tele Form (OpenText, Waterloo, Canada) system which were then scanned and data queries resolved by examining the original forms. The data were stored in a SQL database and extracted as CSV files for later exported to STATA version 17 for analysis.

### Statistical analysis

Descriptive analyses were performed to summarise participant demographic and clinical characteristics, EQ5D dimensions and EQ5D VAS scores reporting the following: absolute values, percentages for categorical variables and median (interquartile ranges) for continuous variables.

Utility score calculations, EQ-5D-3L scores were converted into a health utility score using Zimbabwe’s value (13) set. Value sets are population specific weights that reflect how individuals in a particular country or region value different health states. These are used for converting EQ-5D responses into a single summary score (the utility value). Since no value set is currently available for Malawi, we used the Zimbabwean EQ-5D-3L value set, which is the only one available from SSA with similar context to Malawi.

Health utility scores of one reflects perfect health, zero reflects a state equivalent to death and less than zero is a state considered to be worse than death. Medians and interquartile ranges were reported for utility scores, while means and 95% confidence intervals are presented in the **S2 Supplementary results (S2 Table 1)**

To assess factors associated with HRQoL among sepsis survivors, we conducted separate generalised multivariable linear mixed-effects regression models for EQ-5D utility scores and VAS scores. We used the identity link function on a logit-transformed scale because EQ-5D utility scores include negative values, and VAS score ranged up to 100, so we first rescaled all scores to fall strictly within the (0, 1) interval using a min-max transformation then a normal (Gaussian) distribution was assumed for the logit-transformed outcomes. These models accounted for repeated measures across multiple time points while considering individual variability using random effects at the patient level therefore models included a random intercept for each participant to account for repeated measures over time. Independent variables included demographic factors (age, sex, education, employment status), clinical factors (days unwell, HIV status, active TB status). We included days of follow-up (time from day of recruitment to the last assessment available for each participant) as a covariate to adjust for differences in the timing of data collection and hence variation in follow up duration across individuals. This allowed us to control for potential confounding due to variable follow-up duration, while focusing on the effect of other predictors.

This model estimated average associations between predictors and HRQoL outcomes. To address missing data, we included only participants with at least one follow-up assessment. Given the varying degrees of missingness across time points, mixed models were chosen to maximize available data without requiring complete cases at all time points. To aid interpretation of model results, we considered the minimal important difference (MID) for EQ-5D utility and EQ-VAS scores, which reflects the smallest change in score that is perceived as important(25). For EQ-5D utility values, we referenced previously used MID thresholds ranging from 0.03 to 0.05 and for EQ-VAS, a difference of 7 to 10 points on the 0–100 scale (26,27). These thresholds were used to interpret whether statistically significant findings also reached a magnitude likely to be relevant for clinicians and patient outcomes. In a subsequent analysis, we aimed to explore whether baseline HRQoL could predict recovery trajectory over time among sepsis survivors. We included an interaction term between baseline EQ-5D utility score and days of follow-up into the mixed-effects model. This allowed us to assess whether the rate of change in HRQoL varied by initial EQ-5D utility scores, thereby identifying individuals at risk of persistently poor HRQoL outcomes. This model focused on the predictive value of baseline utility while adjusting for relevant covariates and accounting for within subject correlation over repeated timepoints.

Additionally, we conducted separate multivariable linear regression analyses to explore factors associated with HRQoL at baseline and final assessment. Each analysis was performed independently for EQ-5D utility scores and VAS scores. As part of sensitivity analyses, we re-estimated mixed effect model and baseline multivariable linear regression models for EQ-5D utility and VAS scores, incorporating the Universal Vital Assessment (UVA) score to account for illness severity among sepsis participants. HIV status was excluded from these models to avoid redundancy, as it is a component of the UVA score. This analysis aimed to examine whether baseline illness severity confounded the observed associations with HRQoL.

To compare differences of demographic and clinical characteristics, utility scores and VAS scores among groups, the chi-square test was used for categorical variables. For continuous, non-normal distribution variables, comparisons are done using the Kruskal-Wallis Test and the Mann-Whitney U test to compare between 3 and 2 groups, respectively. A p-value < 0.05 was considered statistically significant. In **S2 Supplementary results (S2 Table 1)** we also provide mean comparisons for EQ5d Utility score and VAS score variables; comparisons are done using one-way ANOVA and independent t-test. Complete case analysis was done using participants with available data for variables included for each specific analysis. All analysis was done using Stata version 18(28) and in **(S2 Table 2)** we summarise EQ5D missing data.

### Ethical approvals

The cohort study was approved by the University of Malawi College of Medicine research ethics committee (approval number P.11/16/2063) and the Liverpool School of Tropical Medicine (approval number 16-062). EuroQol gave an authorization with registration number 74197 to use the Malawi (Chichewa) EQ-5D-3L Paper Self-Complete Version EuroQol Group, Netherlands.

## Results

### Participant demographics and clinical characteristics

The study included 225 sepsis cases, 100 hospital controls, and 100 community controls. **Table 2** summarizes participant demographics and clinical characteristics. The median age was 35.9 years (IQR: 27.8–43.5) in sepsis cases, 40.4 years (IQR: 29.1–48.4) in hospital controls, and 32.5 years (IQR: 23.9–38.4) in community controls (p < 0.001). Among sepsis cases, 143 (63.6%) were HIV-positive, compared to 12 (12.0%) in hospital controls and 18 (18.0%) in community controls, and HIV status was unknown for 60 (60.0%) of community controls (p < 0.001). Tuberculosis (TB) was reported in 37 (16.4%) of sepsis cases, 5 (5.0%) of hospital controls, and 4 (4.0%) of community controls (p < 0.001). 180 (80%) of sepsis cases where severely ill (UVA) and at 180 days of follow-up, 62 (27.6%) had died.

**Table 2:**
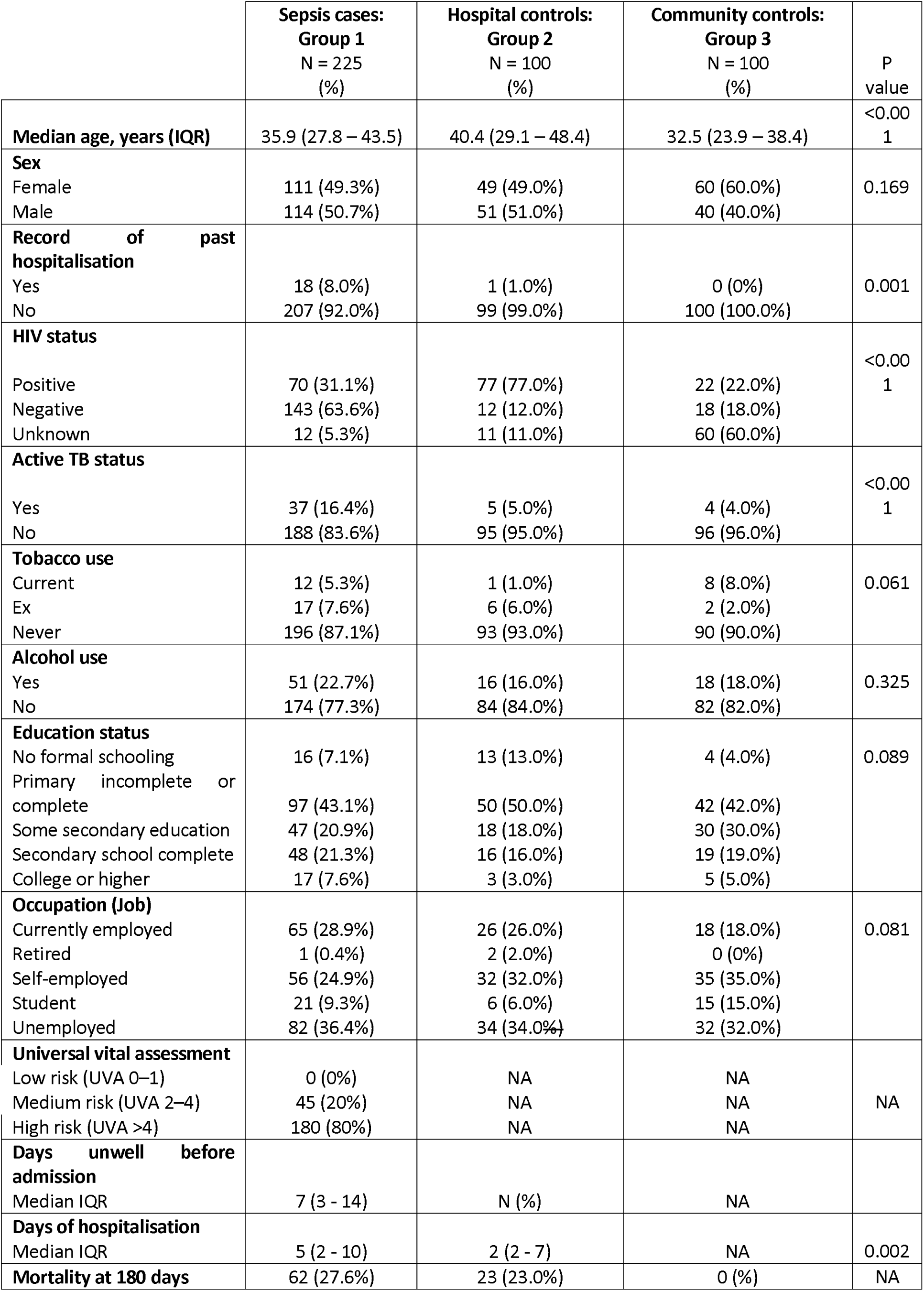
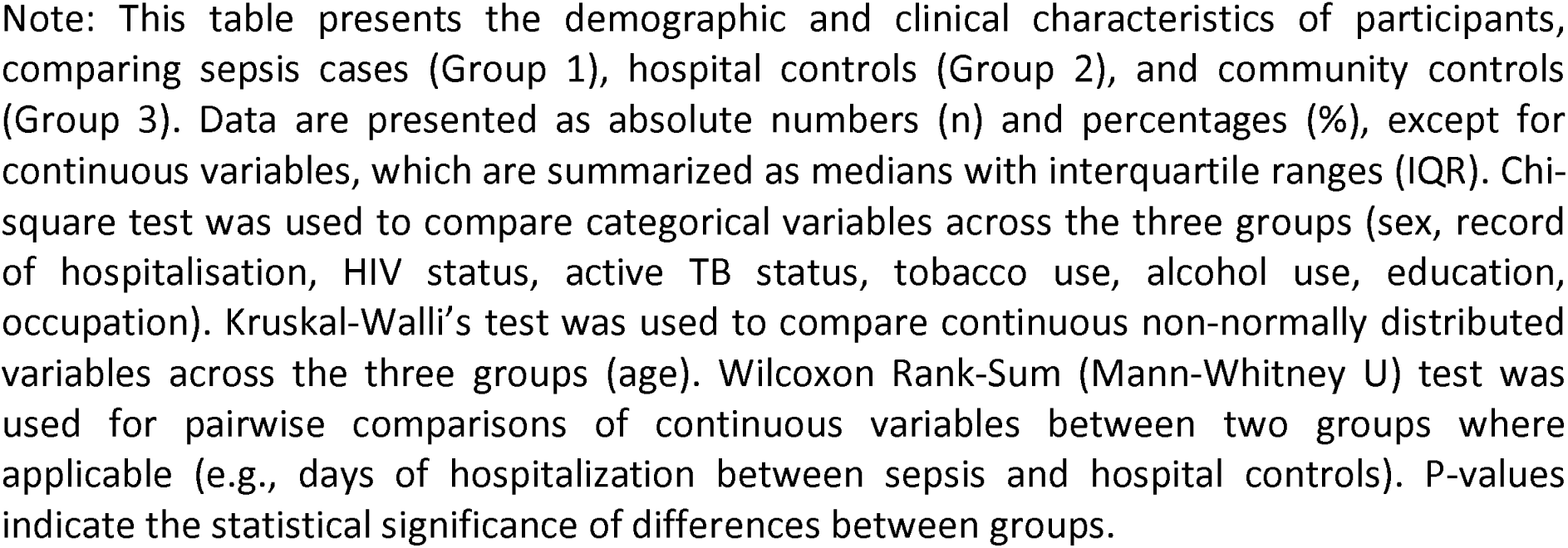
Participants’ demographics and clinical characteristics.

|  | **Sepsis cases:**<br>**Group 1**<br>**N = 225**<br>**(%)** | **Hospital controls:**<br>**Group 2**<br>**N = 100**<br>**(%)** | **Community controls:**<br>**Group 3**<br>**N = 100**<br>**(%)** | **P**<br>**value** |
| --- | --- | --- | --- | --- |
| **Median age, years (IQR)** | 35.9 (27.8 – 43.5) | 40.4 (29.1 – 48.4) | 32.5 (23.9 – 38.4) | <0.001 |
| **Sex** |  |  |  |  |
| Female | 111 (49.3%) | 49 (49.0%) | 60 (60.0%) | 0.169 |
| Male | 114 (50.7%) | 51 (51.0%) | 40 (40.0%) |  |
| **Record of past hospitalisation** |  |  |  |  |
| Yes | 18 (8.0%) | 1 (1.0%) | 0 (0%) | 0.001 |
| No | 207 (92.0%) | 99 (99.0%) | 100 (100.0%) |  |
| **HIV status** |  |  |  |  |
| Positive | 70 (31.1%) | 77 (77.0%) | 22 (22.0%) | <0.001 |
| Negative | 143 (63.6%) | 12 (12.0%) | 18 (18.0%) |  |
| Unknown | 12 (5.3%) | 11 (11.0%) | 60 (60.0%) |  |
| **Active TB status** |  |  |  |  |
| Yes | 37 (16.4%) | 5 (5.0%) | 4 (4.0%) | <0.001 |
| No | 188 (83.6%) | 95 (95.0%) | 96 (96.0%) |  |
| **Tobacco use** |  |  |  |  |
| Current | 12 (5.3%) | 1 (1.0%) | 8 (8.0%) | 0.061 |
| Ex | 17 (7.6%) | 6 (6.0%) | 2 (2.0%) |  |
| Never | 196 (87.1%) | 93 (93.0%) | 90 (90.0%) |  |
| **Alcohol use** |  |  |  |  |
| Yes | 51 (22.7%) | 16 (16.0%) | 18 (18.0%) | 0.325 |
| No | 174 (77.3%) | 84 (84.0%) | 82 (82.0%) |  |
| **Education status** |  |  |  |  |
| No formal schooling | 16 (7.1%) | 13 (13.0%) | 4 (4.0%) | 0.089 |
| Primary incomplete or complete | 97 (43.1%) | 50 (50.0%) | 42 (42.0%) |  |
| Some secondary education | 47 (20.9%) | 18 (18.0%) | 30 (30.0%) |  |
| Secondary school complete | 48 (21.3%) | 16 (16.0%) | 19 (19.0%) |  |
| College or higher | 17 (7.6%) | 3 (3.0%) | 5 (5.0%) |  |
| **Occupation (Job)** |  |  |  |  |
| Currently employed | 65 (28.9%) | 26 (26.0%) | 18 (18.0%) | 0.081 |
| Retired | 1 (0.4%) | 2 (2.0%) | 0 (0%) |  |
| Self-employed | 56 (24.9%) | 32 (32.0%) | 35 (35.0%) |  |
| Student | 21 (9.3%) | 6 (6.0%) | 15 (15.0%) |  |
| Unemployed | 82 (36.4%) | 34 (34.0%) | 32 (32.0%) |  |
| **Universal vital assessment** |  |  |  |  |
| Low risk (UVA 0–1) | 0 (0%) | NA | NA | NA |
| Medium risk (UVA 2–4) | 45 (20%) | NA | NA |  |
| High risk (UVA >4) | 180 (80%) | NA | NA |  |
| **Days unwell before admission** |  |  |  |  |
| Median IQR | 7 (3 - 14) | N (%) | NA |  |
| **Days of hospitalisation** |  |  |  |  |
| Median IQR | 5 (2 - 10) | 2 (2 - 7) | NA | 0.002 |
| **Mortality at 180 days** | 62 (27.6%) | 23 (23.0%) | 0 (%) | NA |

Note: This table presents the demographic and clinical characteristics of participants, comparing sepsis cases (Group 1), hospital controls (Group 2), and community controls (Group 3). Data are presented as absolute numbers (n) and percentages (%), except for continuous variables, which are summarized as medians with interquartile ranges (IQR). Chi-square test was used to compare categorical variables across the three groups (sex, record of hospitalisation, HIV status, active TB status, tobacco use, alcohol use, education, occupation). Kruskal-Wallis's test was used to compare continuous non-normally distributed variables across the three groups (age). Wilcoxon Rank-Sum (Mann-Whitney U) test was used for pairwise comparisons of continuous variables between two groups where applicable (e.g., days of hospitalization between sepsis and hospital controls). P-values indicate the statistical significance of differences between groups.

### EQ-5D-3L dimensions proportions and trends

**Table 3** summarises EQ-5D dimensions across assessments, baseline (day 0), day 7, day 28, day 90, and day 180. At baseline, mobility impairment was reported in 138 (68.6%) of sepsis cases, 43 (53.0%) of hospital controls, and 11 (12.1%) of community controls (p < 0.001). Self-care difficulties were present in 117 (58.0%) of sepsis cases, 25 (30.5%) of hospital controls, and 1 (1.1%) of community controls (p < 0.001). Usual activity limitations were reported in 142 (72.0%) of sepsis cases, 54 (65.0%) of hospital controls, and 4 (4.4%) of community controls (p < 0.001). Pain and discomfort were noted in 138 (71.5%) of sepsis cases, 42 (56.8%) of hospital controls, and 14 (15.1%) of community controls (p < 0.001). Anxiety and depression were reported in 83 (43.2%) of sepsis cases, 34 (43.6%) of hospital controls, and 24 (26.1%) of community controls (p = 0.002).

**Table 3:** EQ-5D-3L Health dimensions across groups at baseline and follow-up assessments.

|  |  | Sepsis cases |  |  |  | Inpatient control |  |  |  | Community controls |  |  | P values |
| --- | --- | --- | --- | --- | --- | --- | --- | --- | --- | --- | --- | --- | --- |
| Baseline | N | No problems | Moderate/some problems | Unable/extreme problems | N | No problems | Moderate/some problems | Unable/extreme problems | N | No problems | Moderate/some problems | Unable/extreme problems |  |
| Mobility | 201 | 63 (31.3%) | 105 (52.2%) | 33 (16.4%) | 81 | 38 (46.9%) | 34 (42.0%) | 9 (11.1%) | 91 | 80 (87.9%) | 11 (12.1%) | 0 (0%) | 0.000 |
| Selfcare | 202 | 85 (42.1%) | 79 (39.1%) | 38 (18.8%) | 82 | 57 (69.5%) | 16 (19.5%) | 9 (11.0%) | 92 | 91 (98.9%) | 1 (1.1%) | 0 (0%) | 0.000 |
| Usual activity | 197 | 55 (27.9%) | 84 (42.6%) | 58 (29.4%) | 83 | 29 (34.9%) | 32 (38.6%) | 22 (26.5%) | 92 | 88 (95.7%) | 4 (4.4%) | 0 (0%) | 0.000 |
| Pain and discomfort | 193 | 55 (28.5%) | 99 (51.3%) | 39 (20.2%) | 74 | 32 (43.2%) | 33 (44.6%) | 9 (12.2%) | 93 | 79 (85.0%) | 14 (15.1%) | 0 (0%) | 0.000 |
| Anxiety and depression | 192 | 109 (56.8%) | 77 (40.1%) | 6 (3.1%) | 78 | 44 (56.4%) | 26 (33.3%) | 8 (10.3%) | 92 | 68 (73.9%) | 24 (26.1%) | 0 (0%) | 0.002 |
| Day 7 |  |  |  |  |  |  |  |  |  |  |  |  |  |
| Mobility | 153 | 74 (48.4%) | 63 (41.2%) | 16 (10.5%) | 54 | 34 (63.0%) | 17 (31.5%) | 3 (5.6%) | NA | NA | NA | NA | 0.910 |
| Selfcare | 150 | 82 (54.7%) | 54 (36.0%) | 14 (9.3%) | 51 | 39 (76.5%) | 9 (17.7%) | 3 (5.9%) | NA | NA | NA | NA | 0.221 |
| Usual activity | 145 | 56 (38.6%) | 57 (39.3%) | 32 (22.1%) | 52 | 18 (34.6%) | 26 (50.0%) | 8 (15.4%) | NA | NA | NA | NA | 0.756 |
| Pain and discomfort | 151 | 62 (41.1%) | 72 (47.7%) | 17 (11.3%) | 51 | 29 (56.9%) | 21 (41.2%) | 1 (2.0%) | NA | NA | NA | NA | 0.685 |
| Anxiety and depression | 147 | 84 (57.1%) | 54 (36.7%) | 9 (6.1%) | 52 | 32 (61.5%) | 18 (34.6%) | 2 (3.9%) | NA | NA | NA | NA | 1.000 |
| Day 28 |  |  |  |  |  |  |  |  |  |  |  |  |  |
| Mobility | 106 | 67 (63.2%) | 35 (33.0%) | 4 (3.8%) | 30 | 22 (73.3%) | 8 (26.7%) | 0 (0%) | 56 | 52 (92.9%) | 4 (7.1%) | 0 (0%) | 0.172 |
| Selfcare | 105 | 77 (73.3%) | 25 (23.8%) | 3 (2.9%) | 30 | 25 (83.3%) | 5 (16.7%) | 0 (0%) | 57 | 57 (100.0%) | 0 (0%) | 0 (0%) | 0.089 |
| Usual activity | 106 | 57 (53.7%) | 32 (30.2%) | 17 (16.0%) | 28 | 18 (64.3%) | 9 (32.1%) | 1 (3.6%) | 58 | 55 (94.8%) | 3 (5.2%) | 0 (0%) | 0.251 |
| Pain and discomfort | 107 | 55 (51.4%) | 47 (43.9%) | 5 (4.7%) | 30 | 16 (53.3%) | 13 (43.3%) | 1 (3.3%) | 57 | 46 (80.7%) | 11 (19.3%) | 0 (0%) | 0.086 |
| Anxiety and depression | 103 | 63 (61.2%) | 35 (34.0%) | 5 (4.9%) | 28 | 23 (82.1%) | 5 (17.9%) | 0 (0%) | 58 | 46 (79.3%) | 12 (20.7%) | 0 (0%) | 0.506 |
| Day 90 |  |  |  |  |  |  |  |  |  |  |  |  |  |
| Mobility | 56 | 40 (71.4%) | 15 (26.8%) | 1 (1.8%) | 14 | 9 (64.3%) | 5 (35.7%) | 0 (0%) | NA | NA | NA | NA | 1.000 |
| Selfcare | 59 | 48 (81.4%) | 10 (17.0%) | 1 (1.7%) | 14 | 11 (78.6%) | 3 (21.4%) | 0 (0%) | NA | NA | NA | NA | 0.913 |
| Usual activity | 59 | 43 (72.9%) | 11 (18.6%) | 5 (8.5%) | 15 | 9 (60.0%) | 5 (33.3%) | 1 (6.7%) | NA | NA | NA | NA | 1.000 |
| Pain and discomfort | 60 | 33 (55.0%) | 26 (43.3%) | 1 (1.7%) | 13 | 5 (38.5%) | 8 (61.5%) | 0 (0%) | NA | NA | NA | NA | 1.000 |
| Anxiety and depression | 60 | 45 (75.0%) | 13 (21.7%) | 2 (3.3%) | 15 | 8 (53.3%) | 7 (46.7%) | 0 (0%) | NA | NA | NA | NA | 0.508 |
| Day 180 |  |  |  |  |  |  |  |  |  |  |  |  |  |
| Mobility | 26 | 17 (65.4%) | 9 (34.6%) | 0 (0%) | 10 | 8 (80.0%) | 2 (20.0%) | 0 (0%) | 25 | 22 (88.0%) | 3 (12.0%) | 0 (0%) | 0.670 |
| Selfcare | 25 | 21 (84.0%) | 3 (12.0%) | 1 (4.0%) | 10 | 10 (100%) | 0 (0%) | 0 (0%) | 25 | 24 (96.0%) | 1 (4.0%) | 0 (0%) | 0.670 |
| Usual activity | 26 | 18 (69.2%) | 6 (23.1%) | 2 (7.7%) | 11 | 8 (72.7%) | 3 (27.3%) | 0 (0%) | 25 | 23 (92.0%) | 2 (8.0%) | 0 (0%) | '0.670 |
| Pain and discomfort | 24 | 18 (75.0%) | 6 (25.0%) | 0 (0%) | 9 | 7 (77.8%) | 2 (22.2%) | 0 (0%) | 26 | 19 (73.1%) | 7 (26.9%) | 0 (0%) | 0.419 |
| Anxiety and depression | 26 | 21 (80.8%) | 4 (15.4%) | 1 (3.9%) | 10 | 7 (70.0%) | 2 (20.0%) | 1 (10.0%) | 26 | 21 (80.8%) | 5 (19.2%) | 0 (0%) | '0.670 |

Note: This table presents comparisons of ordinal EQ-5D-3L health dimensions (Mobility, Self-care, Usual Activities, Pain/Discomfort, and Anxiety/Depression) across assessment time points (Baseline, day 7, day 28, day 90, and day 180) among sepsis cases, hospital controls, and community controls. Data are presented as frequency counts (N) and percentages (%) for each EQ-5D-3L dimension level (No problems, Moderate/Some problems, Unable/Extreme problems). Kruskal-Wallis (KW) test was used to compare ordinal EQ-5D-3L dimensions across all three groups (sepsis cases, hospital controls, and community controls). Wilcoxon Rank-Sum (Mann-Whitney U) test was used for pairwise comparisons between two groups (sepsis cases vs. hospital controls). P-values indicate the statistical significance of differences between groups at each time point.

Figure 1 shows the trend for proportions of participants reporting no problems, moderate/some problems, and unable/extreme problems across five EQ-5D-3L dimensions at different time points (baseline, day 7, day 28, day 90, and day 180). Sepsis cases had the highest proportion of impairments, with gradual improvement over time, but persistent limitations remained at day 180. Hospital controls showed similar trends but with fewer reported problems, while community controls consistently reported minimal health issues throughout the study. Across all assessments, usual activity remained the most affected domain among sepsis cases, with limitations persisting up to day 180. Anxiety and depression were the next most frequently reported impairments. While mobility, self-care, and usual activities improved over time, some residual disability persisted at day 180.

**Fig 1:**
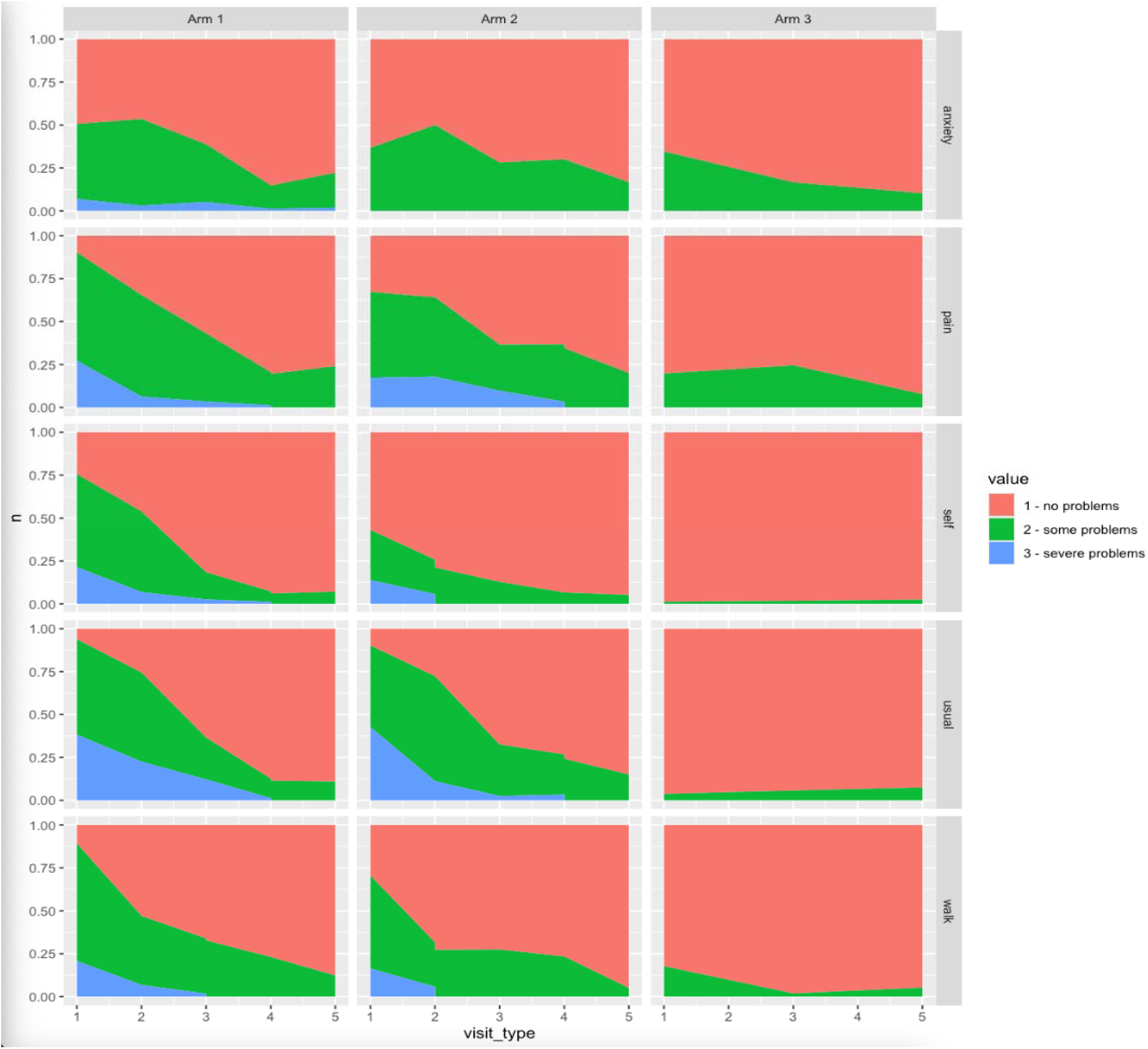
Proportions of participants with no problems, moderate/some problems and unable/extreme problems among groups across assessment. Note: A stacked area plot utility scores and visual analogue scale scores (VAS scores). Arm1 = Sepsis cases, Arm 2= Inpatient controls. Arm 3 = Community controls

**Table 4** summarises median utility scores and VAS scores across groups and assessments. At baseline, median utility scores were 0.596 (IQR: 0.365–0.734) in sepsis cases, 0.666 (IQR: 0.611–0.722) in hospital controls, and 0.900 (IQR: 0.833–0.900) in community controls. By day 180, the median utility score for sepsis cases declined to 0 (IQR: 0–0), while hospital controls improved to 0.900 (IQR: 0.900–0.900).

**Table 4:** Participants’ summary of utility scores and VAS scores.

|  | **Sepsis cases**<br>**(including deceased)** | **Hospital control** | **Community controls** | **P value** |
| --- | --- | --- | --- | --- |
|  | [N]Median (IQR) | [N]Median (IQR) | [N]Median (IQR) |  |
| **Baseline Utility score** | [189]<br>0.596 (0.365 - 0.734) | [76]<br>0.666 (0.611 - 0.722) | [91]<br>0.900 (0.833 - 0.900) | <0.001 |
| **Day 7 Utility score** | [134]<br>0.790 (0.642- 0.900) | [40]<br>0.789 (0.688 - 0.900) | NA | 0.661 |
| **Day 28 Utility score** | [124]<br>0.642 (0 - 0.900) | [21]<br>0.844 (0.734 - 0.900) | [53]<br>0.9 (0.854 - 0.900) | <0.001 |
| **Day 90Utility score** | [88]<br>0 (0 - 0.839) | [13]<br>0.9 (0.833 - 0.900) | NA | <0.001 |
| **Day 180 Utility score** | [71]<br>0 (0 - 0) | [2]<br>0.9 (0.900 - 0.900) | [20]<br>0.900 (0.900 - 0.900) | <0.001 |
| **Baseline VAS score** | [210]<br>62.5 (50 - 80) | [87]<br>65 (45 - 80) | [93]<br>90 (80 - 90) | <0.001 |
| **Day 7 VAS score** | [157]<br>70 (60 - 85) | [55]<br>75 (60 - 90) | NA | 0.348 |
| **Day 28 VAS score** | [107]<br>80 (70 - 90) | [31]<br>80 (70 - 90) | [59]<br>90 (85 - 90) | <0.001 |
| **Day 90 VAS score** | [61]<br>85 (70 - 90) | [15]<br>80 (70 - 90) | NA | 0.217 |
| **Day 180 VAS score** | [27]<br>90 (70 - 95) | [11]<br>90 (75 - 95) | [26]<br>90 (80 - 95) | 0.985 |

Note: Data are presented as median (interquartile range, IQR) with the number of participants [N] in each group. Kruskal-Wallis's test was used to compare utility and VAS scores across all three groups (sepsis cases, hospital controls, and community controls) at each time point. Mann-Whitney U test was used for pairwise comparisons between two groups (sepsis cases vs. hospital controls). p-values indicate the statistical significance of differences between groups. NA (Not Applicable) indicates time points where data was not collected for the group.

**S2 Table 1** presents mean utility and VAS scores across assessments. Mean baseline utility scores were 0.539 (95% CI: 0.500–0.578) in sepsis cases, 0.666 (95% CI: 0.611–0.722) in hospital controls, and 0.861 (95% CI: 0.850–0.874) in community controls. By day 180, mean utility scores declined to 0.133 (95% CI: 0.059–0.208) in sepsis cases, while hospital controls improved to 0.900 (95% CI: 0.900–0.900). Utility scores declined most in sepsis cases, while hospital and community controls remained stable (**S2 Figure 1**). VAS scores improved over time for sepsis cases (**S2 Figure 2**).

Figure 2 presents box plots of EQ-5D utility scores across five time points (day 0, day 7, day 28, day 90, and day 180) for sepsis cases, hospital controls, and community controls. Utility scores improved for day 7 and day 28, then declined in day 90 and day 180 assessments in sepsis cases, while hospital and community controls showed stable or improving trends. VAS scores increased over time for sepsis cases, hospital controls maintain relatively high VAS scores, and community controls consistently report the highest VAS scores (Figure 3).

**Fig 2:**
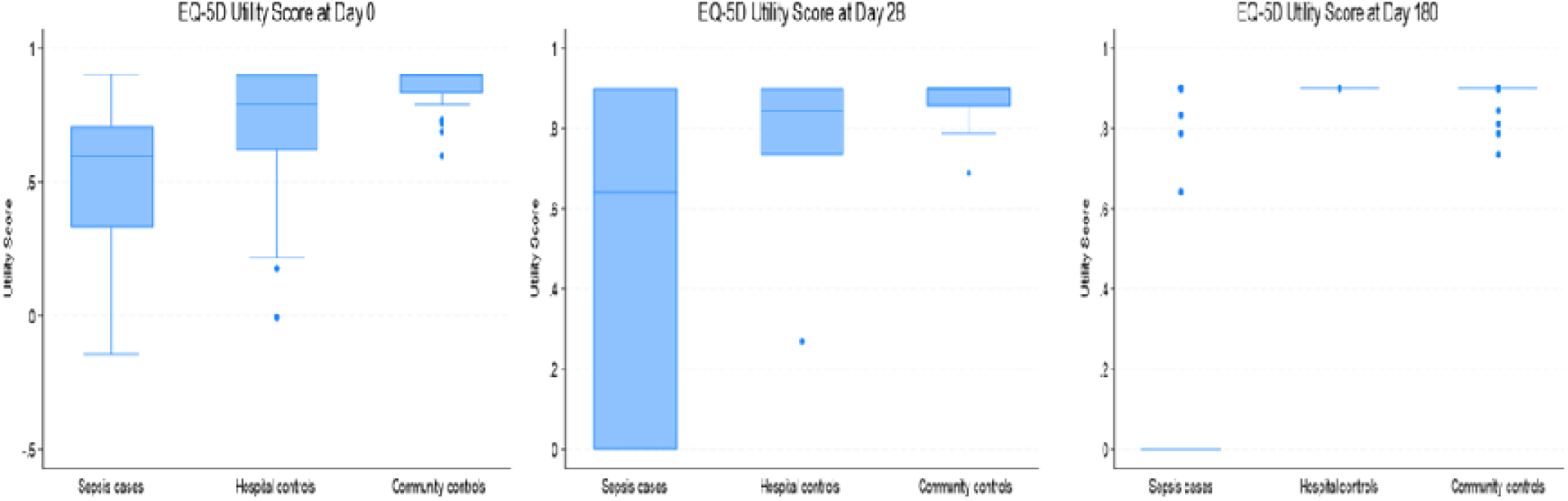
Longitudinal trends in EQ-5D utility scores across 180 days in sepsis cases, hospital controls, and community controls. Note: Boxplots show the distribution of EQ-5D utility scores for participants in each study group on Day 0, Day 28 and Day 180. The horizontal line within each box represents the median, while the box edges indicate the interquartile range (IQR). Whiskers extend to the lowest and highest values within 1.5 × IQR; values outside this range are plotted as outliers.

**Fig 3:**
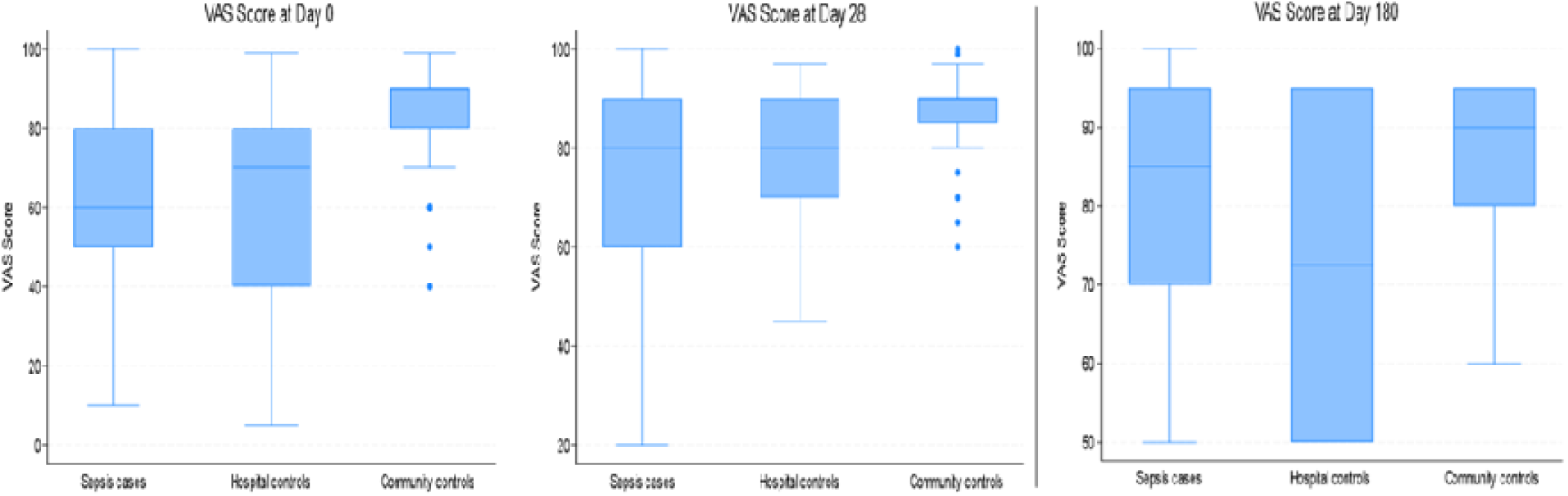
Longitudinal trends in EQ-5D VAS scores across 180 days in sepsis cases, hospital controls, and community controls. Note: Boxplots show the distribution of VAS scores for participants in each study group on Day 0, Day 28, and Day 180. The horizontal line within each box represents the median, while the box edges indicate the interquartile range (IQR). Whiskers extend to the lowest and highest values within 1.5 × IQR; values outside this range are plotted as outliers.

### Factors associated with HRQoL in sepsis patients

In the multivariable mixed effects regression models to examine HRQoL among sepsis cases at multiple time points Table 5, Increased duration of illness prior to admission was significantly associated with lower HRQoL, with each additional day unwell predicting a small but statistically significant decline in logit-transformed EQ-5D scores (β = -0.006, p < 0.004) .Additionally, participants with unknown HIV status reported significantly higher EQ-VAS scores (β = 1.15, p = 0.023). Sepsis survivors who report higher HRQoL at baseline are more likely to continue reporting better HRQoL across follow-up (β = 0.465, p < 0.001). Associations persisted even with inclusion of UVA in the model.

**Table 5.** Socio-demographic and clinical factors associated with HRQoL for sepsis cases (multivariable mixed effect regression model)

|  | Utility score<br>β (95% CI)<br>P value<br>N=606 | Utility score + UVA<br>β (95% CI)<br>P value<br>N=545 | VAS score<br>β (95% CI)<br>P value<br>N=562 | VAS score + UVA β<br>(95% CI)<br>P value<br>N=504 |
| --- | --- | --- | --- | --- |
| **Age (years)** | -0.012 (-0.035, 0.011);<br>p=0.293 | -0.015 ( -0.041,<br>0.010); p=0.252 | -0.000 (-0.019,<br>0.020); p=0.987 | -0.007 ( -0.026,<br>0.012); p=0.493 |
| **Sex (Ref female)** |  |  |  |  |
| Male | 0.105 (-0.442, 0.651);<br>p=0.707 | 0.186 ( -0.383,<br>0.754); p= 0.522 | 0.278 (-0.179,<br>0.736); p=0.233 | 0.310 ( -0.120,<br>0.739); p=0.158 |
| **HIV status (Ref negative)** |  |  |  |  |
| Positive | -0.244 (-0.799, 0.311);<br>p=0.390 | - | -0.020 (-0.480,<br>0.438); p= 0.929 | - |
| Unknown | 0.619 (-0.509, 1.746);<br>p=0.282 | - | 1.154 (0.162,<br>2.145); p=0.023 | - |
| **Active TB status (Ref no)** |  |  |  |  |
| Yes | -0.162 (-0.789 0.464);<br>p=0.611 | -0.123 ( -0.747,<br>0.501); p= 0.699 | -0.056 ( -0.595,<br>0.482); p=0.838 | -0.035 (-0.524,<br>0.454); p=0.889 |
| **Education status (Ref college or higher)** |  |  |  |  |
| No formal schooling | -0.310 (-1.541, 0.920);<br>p=0.621 | -0.505 ( -1.884,<br>0.874); p= 0.473 | 0.039 (-1.090,<br>1.167); p=0.946 | 0.028 (-1.124,<br>1.179); p=0.963 |
| Primary incomplete or complete | -0.080 (-0.991, 0.832);<br>p=0.864 | -0.107 (-1.049,<br>0.834); p=0.823 | 0.146 (-0.647,<br>0.938); p=0.719 | -0.022 ( -0.777,<br>0.732); p=0.954 |
| Secondary school complete | 0.119 (-0.848, 1.086);<br>p=0.809 | -0.071 (-1.066,<br>0.924); p=0.890 | 0.190 (-0.640,<br>1.019); p= 0.654 | -0.031 ( -0.822,<br>0.760); p=0.939 |
| Some secondary education | 0.197 (-0.766, 1.160);<br>p=0.689 | -0.091 (-1.109,<br>0.928); p=0.862 | 0.547 (-0.270,<br>1.364); p=0.190 | 0.104 (-0.690,<br>0.898); p=0.798 |
| **Occupation (Ref currently employed)** |  |  |  |  |
| Retired | 3.303 (-0.553, 7.160);<br>p=0.093 | 3.562 ( -0.297,<br>7.421); p=0.070 | -0.328 (-3.038,<br>2.382); p=0.937 | -0.306 (-2.77, 2.162);<br>p=0.808 |
| Self-employed | -0.137 (-0.826, 0.550);<br>p=0.695 | 0.091 (-0.628,<br>0.811); p=0.803 | -0.330 (-0.911,<br>0.251); p=0.265 | -0.033 (-0.584,<br>0.519); p=0.908 |
| Student | 0.125 (-0.862, 1.113);<br>p=0.803 | 0.186 (-0.869,<br>1.241); p=0.729 | -0.417 (-1.227,<br>0.392); p=0.312 | -0.333 (-1.124,<br>0.458); p=0.410 |
| Unemployed | 0.008 (-0.711, 0.695);<br>p=0.981 | -0.009 ( -0.733,<br>0.714); p=0.980 | -0.394 (-0.985,<br>0.196); p=0.191 | -0.337 ( -0.889,<br>0.214); p=0.230 |
| **Days unwell before admission** | -0.006 (-0.010, -0.002);<br>p=0.004 | -0.009 ( -0.018, -<br>0.000); p= 0.046 | -0.003 (-0.006,<br>0.001); p=0.102 | -0.004 (-0.012,<br>0.003); p=0.234 |
| **UVA** | - | -0.095 ( -0.254,<br>0.064); p= 0.243 | - | 0.023 (-0.102,<br>0.149); p= 0.715 |
| **Baseline HRQoL score** | 0.465 (0.339, 0.591);<br>p<0.001 | 0.440 (0.305,<br>0.576); p<0.001 | 0.813 (0.606,<br>1.022); p<0.001 | 0.826 (0.612, 1.041);<br>p<0.001 |

Note: This table presents the results of the linear mixed-effects regression model, showing the coefficients (95% Confidence Intervals, CI) and p-values for factors associated with EQ-5D utility scores and VAS scores. Random effects were included at the patient level to account for within-subject correlation across time. A negative coefficient indicates a decrease in utility or VAS score associated with that variable. A positive coefficient indicates an increase in utility or VAS score associated with that variable.

## Discussion

This study provides critical insights into the long-term HRQoL outcomes among sepsis survivors in Malawi. Sepsis survivors in Malawi experience significant and prolonged impairments in quality of life, which persist well beyond hospital discharge. These findings reinforce existing evidence that sepsis survivors experience prolonged physical and mental health challenges (29).The impact of sepsis extends long after the acute phase and may influence survivors’ ability to live independently and participate fully in daily life (14). Further research work is required to explore tailored context specific interventions that address both physical and mental health outcomes in sepsis survivors, particularly in low-resource settings. We recommend that future sepsis interventions and research prioritize not only survival but also long-term quality of life as a key outcome measure (30).

Our study found that sepsis leads to a significant and sustained deterioration in HRQoL across physical, psychological, and functional domains. Across all EQ-5D dimensions, sepsis survivors reported persistent impairments through 180 days, with mobility, pain/discomfort, and usual activities most affected. Compared to hospital and community controls, sepsis survivors had markedly lower utility scores from baseline to day 180, highlighting a disproportionate and prolonged burden. While EQ-5D VAS scores improved modestly, utility scores declined over time, reflecting a disconnect between perceived and actual functional recovery. We also found that delays in hospital presentation and lower baseline EQ-5D utility scores were significantly associated with worse post-sepsis HRQoL. Older age and female sex predicted lower scores, aligning with broader literature on post-acute outcomes.

Our findings are consistent with other studies in demonstrating long-term physical and psychological impairments in sepsis survivors. On physical impact, Yende et al. (2016) reported that one-third of previously independent sepsis survivors could not return to independent living by six months, with common impairments in mobility (37.4%) and usual activities (43.7%) (31). Like our findings, these impairments persisted or worsened over time showing that physical recovery post-sepsis is slow and often incomplete, especially in resource-limited settings where post-discharge(8,32,33). Over 30% of our participants experienced mobility limitations at six months, similar to other studies which identifying physical disability as one of the most persistent sequelae of sepsis(9,33). In our study, pain and discomfort were also prevalent, aligning with research that associates sepsis with chronic pain and therefore reduced HRQoL (30). Psychological distress including anxiety and depression remained a significant concern, supporting the need for psychosocial interventions. Prior research, such as those involving hepatitis B patients, have shown that strong social support can improve mental health outcomes, suggesting that similar models may benefit sepsis survivors (11). While there is evidence of partial psychological recovery by six months (33), our findings suggest that mental health challenges may be more prolonged in low-resource settings, where access to psychosocial care is often limited (30,34). The findings suggest that sepsis not only impacts individuals but also may affect families and society, through increased caregiving needs, reduced productivity, and ongoing dependency (35,36). For example, in Malawi, hospitalizations for bloodstream infections have been associated with substantial economic impacts, with resistant infections incurring significantly poorer HRQoL(14).

Su et al. (2018) also identified female sex and older age as predictors of poorer QoL factors and thus that mirrored our regression results (37). Furthermore, there has been an association of age and sex to significantly impact HRQoL outcomes, with older age and female sex associated more specifically with worse physical health scores(8,38), in our study, these factors were associated with lower utility and VAS scores. Some studies however argue that sepsis survivors show higher rates of chronic conditions, therefore the impact on HRQoL for older people, may be due to pre-existing trajectories rather than sepsis itself (39). Evidence on elderly populations in low-resource settings have shown that other socioeconomic factors, such as income, education, and social support, significantly influence general QoL, highlighting the need for holistic interventions on elderly people (40). Overall, sepsis has substantial long-term impacts on survivors’ quality of life and independence (35,36).

In both EQ-5D utility and EQ-VAS models, we identified factors associated with meaningful differences in health-related quality of life among sepsis survivors. For EQ-5D, a longer duration of illness prior to admission was significantly associated with lower utility scores. Although the per-day effect was modest (β = −0.0061), the cumulative impact over a 30-day period approached a reduction of 0.04 units, which is within the commonly accepted MID range of 0.03–0.05 (26,27). This suggests that delays in care may have a clinically significant effect on post-sepsis HRQoL. Patients with higher baseline EQ-5D scores experienced consistently better quality of life throughout the follow up, suggesting that a simple EQ-5D assessment during admission could be a useful tool for identification of risk individuals and planning targeted follow-up of sepsis survivors with low baseline HRQoL. In the EQ-VAS model, participants with unknown HIV status reported significantly higher self-rated health compared to those with known status. While the predicted difference exceeded the MID threshold of 7–10 points for EQ-VAS this result may reflect selection or reporting bias given the small subgroup size.

EQ-VAS scores showed a trend of improvement over time while EQ-5D utility scores declined. This discrepancy is consistent with prior literature and reflects the conceptual differences between the two measures. EQ-VAS captures patients’ single reading subjective perceptions of health and is therefore sensitive to psychological and emotional adaptation, which may improve during recovery even if functional limitations persist. In contrast, EQ-5D utility scores are derived from predefined multiple health dimensions and anchored to population preferences, which may remain low due to residual impairments such as pain, anxiety, or limited mobility. These findings reinforce the value of using both instruments to capture different aspects of health-related quality of life in post-sepsis populations(41,42) therefore EQ-VAS may reflect psychological adaptation, while EQ-5D captures ongoing functional deficits.

While most global studies describe HRQoL impairments, few have explored sepsis outcomes in sub-Saharan Africa. Our data point to a sustained HRQoL decline in sub-Saharan Africa with a severe and progressive decline in utility scores among sepsis survivors in Malawi by day 180. This contrasts with findings from Granja et al. and Su et al., where HRQoL either remained stable or improved over time and was comparable to other ICU survivors(37,43). This difference may potentially be due to structural and contextual factors. For instance, a high proportion of sepsis survivors in our study had pre-existing HIV or TB infections, conditions known to delay recovery and increase post-sepsis morbidity (7,44). In addition, the lower baseline QoL in SSA populations may contribute to steeper declines after critical illness (45). Furthermore, unlike high-income countries where post-sepsis care pathways, including physiotherapy and mental health services, are well established, such services (46)are limited or absent in Malawi. These systemic gaps likely compound the effects of sepsis on HRQoL and recovery. These findings highlight the urgent need for context-specific interventions to improve long-term outcomes for sepsis survivors in Malawi and SSA at large. As recommended by other studies (47,48) future strategies should integrate rehabilitation programs, although they may not decrease mortality, they may improve quality of life. Given the significant impact of sepsis on survivors’ quality of life, our findings suggest that both mortality and HRQoL should be considered when designing interventions and endpoints for sepsis studies and care. (30).

### Strengths of the study

This study is one of the first in SSA to assess long-term HRQoL outcomes in sepsis survivors, providing valuable regional data. The inclusion of both hospital and community controls strengthens the study’s comparative design. The inclusion of community controls allowed us to show that sepsis survivors start from a lower baseline and diverge further over time, unlike in other studies where HRQoL was benchmarked against other hospital survivors only. Also, most global reports exclude the EQ-VAS, yet we demonstrate its value in capturing subjective recovery distinct from functional change.

### Limitations of the study

This study focused on hospitalised sepsis cases, which may underestimate the burden in community-managed cases and provide selection bias. Mortality and high loss to follow-up may have led to an underestimation of HRQoL recovery over time. However, its generalisation in SSA is limited, and value sets for many African populations, including Malawi, are unavailable. It was necessary to use Zimbabwean value sets due to the lack of Malawi-specific utility weights, and while this approach was commonly used in such contexts, it may introduce some minor estimation error, as preferences for health states may differ between Zimbabwean and Malawian populations. Zimbabwean value set for EQ5D3L is the only one available from SSA, the absence of alternative SSA value sets also limited our ability to conduct sensitivity analyses comparing how the use of different value sets might influence our findings.

While our models accounted for repeated measures over time using random intercepts, we did not include random slopes for time, which would have allowed us to fully characterize individual variation in longitudinal recovery trajectories. We did not perform growth curve modelling (e.g., random slopes and intercepts) or moderation/mediation analyses due to the exploratory nature of this study and our primary focus on identifying key associations rather than establishing causal pathways. Growth curve modelling could have provided insights into both average and individual-specific recovery trajectories, and mediation or moderation analyses could have uncovered potential mechanisms, upstream effect or modifiers that might have affected the outcomes of the models, such advanced modelling was beyond the scope of our current objectives. Furthermore, these models are more complex, require stronger assumptions, and may not be easily interpretable or estimable with limited follow-up data affected by the level of missingness in our data. Future work should consider this approach to better describe heterogeneity in recovery and identify subgroups needing targeted support.

## Conclusion

This study provides evidence of the long-term HRQoL deficits among sepsis survivors in Malawi, with the persistent impact on physical and mental well-being. Our findings highlight the urgent need for post-sepsis rehabilitation strategies, particularly in resource-limited settings like Malawi. By integrating both clinical and quality-of-life measures, we can enhance post-sepsis care and reduce the long-term disease burden in vulnerable populations.

Our findings underscore the urgent need for structured post-sepsis care models in SSA, which could include physiotherapy, mental health support, and pain management services. Post-COVID-19 rehabilitation programs in improving HRQoL suggest that similar approaches could be applied to sepsis survivors in SSA(49). Future research should focus on developing tailored interventions that address pain management, mobility recovery, and mental health support, ultimately improving long-term outcomes for sepsis survivors in SSA. Also, research should focus on developing Malawi-specific EQ-5D-3L value sets to enhance the accuracy of HRQoL assessments and evaluating cost-effective rehabilitation interventions to improve long-term outcomes for sepsis survivors.

## Supporting information

Supplementary results

Study Checklist

## Data Availability

All data underlying the findings of this study are fully available without restriction. The datasets have been deposited in publicly accessible repositories and can be accessed via the following links: https://github.com/joelewis101/blantyreESBL https://github.com/joelewis101/blantyreSepsis

https://github.com/joelewis101/blantyreSepsis

## Acknowledgements

We would like to acknowledge the clinical team at Queen Elizabeth Central Hospital, and the research assistants for making the collection of this data successful. We also acknowledge Sarah White of the international public health department at Liverpool School of Tropical Medicine for her statistical guidance.

## List of captions for supplementary documents

1. S1 Checklist for planning studies including HRQoL measures CHEST 2020; 158(1S): S49-S56
2. S2 Table 1: Mean participants’ utility scores and VAS at different time points
3. S2 Table 2: Summary of HRQoL data missingness at different time points
4. S2 Table 3: Multivariable regression models for EQ-5D utility and VAS scores at baseline and day 180 for sepsis cases
5. S2 Figure 1: Mean EQ-5D utility scores across groups at different time points
6. S2 Figure 2: Mean EQ-5D VAS scores across groups at different time points

