## Supplementary results for "Health-related quality of life among patients with sepsis in Blantyre, Malawi: an observational cohort study"

### S2 Supplementary results

#### S2 Table 1: Mean participants' utility scores and VAS at different time points

|  | **Sepsis cases (including deceased)** | **Hospital control** | **Community controls** | **P value** |
| --- | --- | --- | --- | --- |
|  | [N]Mean (95%CI) | [N]Mean (95%CI) | [N]Mean (95%CI) |  |
| **Baseline utility score** | [189] 0.539 (0.500 - 0.578) | [76] 0.760 (0.561 - 0.856) | [91] 0.861 (0.849 - 0.874) | <0.001 |
| **Day 7 utility score** | [134] 0.739 (0.705 - 0.772) | [40] 0.764 (0.714 - 0.814) | - | 0.453 |
| **Day 28 utility score** | [124] 0.552 (0.482 - 0.622) | [21] 0.823 (0.758 - 0.887) | [53] 0.872 (0.859 - 0.886) | <0.001 |
| **Day 90 utility score** | [88] 0.360 (0.270 - 0.450) | [13] 0.846 (0.791 - 0.902) | - | <0.001 |
| **Day 180 utility score** | [71] 0.133 (0.059 - 0.208) | [2] 0.900 (0.900 - 0.900) | [20] 0.879 (0.857 - 0.901) | <0.001 |
| **Baseline VAS score** | [210] 64.1 (61.4 - 66.8) | [87] 61.4 (56.4 - 66.5) | [93] 85.4 (83.3 - 87.5) | <0.001 |
| **Day 7 VAS score** | [157] 69.6 (66.7 - 72.6) | [55] 71.4 (65.7- 77.0) | - | 0.570 |
| **Day 28 VAS score** | [107] 76.2 (72.8 - 79.5) | [15] 76.0 (68.2 - 83.8) | [59] 87.7 (85.6 - 89.8) | <0.001 |
| **Day 90 VAS score** | [61] 79.4 (75.0 -83.9) | [31] 77.2 (71.8 - 82.5) | - | 0.480 |
| **Day 180 VAS score** | [27] 83.4 (77.5 - 89.3) | [11] 82.6 (70.8 - 94.4) | [26] 86.3 (82.2 -90.3) | 0.669 |

Note: This table presents mean (95% Confidence Interval, CI) Utility Scores and VAS scores across five assessment time points (Baseline [Day 0], Day 7, Day 28, Day 90, and Day 180) among sepsis cases, hospital controls, and community controls. Utility scores range from 0 (death) to 1 (perfect health), derived from the EQ-5D-3L index. VAS scores range from 0 (worst health state) to 100 (best health state), representing self-reported health perception. Data are presented as a means with 95% Confidence Intervals (CI) and number of participants [N] per group. One-way ANOVA was used to compare mean Utility and VAS scores across all three groups at each time point. Independent T-tests were used for pairwise comparisons between two groups at each time point (sepsis vs. hospital controls). p-values indicate the statistical significance of differences between groups at each time point. NA (Not Applicable) indicates assessments where nor done for the groups

#### S2 Table 2: Summary of HRQoL data missingness at different time points

| **Variable** | **n (%) of missingness**  **N=425** |
| --- | --- |
| Utility score at baseline | 19 (4.5%) |
| Utility score day 7 | 207 (48.7%) |
| Utility score day 28 | 130 (30.6%) |
| Utility scores day 90 | 227 (53.4%) |
| Utility scores day 180 | 198 (46.6%) |
| VAS score at baseline | 12 (2.8%) |
| VAS score day 7 | 171 (40.2%) |
| VAS score day 28 | 155 (36.5%) |
| VAS scores day 90 | 258 (60.7%) |
| VAS scores day 180 | 249 (58.6%) |

#### S2 Table 3: Multivariable regression models for EQ-5D utility and VAS scores at baseline and day 180 for sepsis cases

| **Variable** | **Utility score (Baseline)**  **N=189**  **β (95% CI)**  **p value** | **Utility score (Baseline + UVA)**  **N=170**  **β (95% CI)**  **p value** | **Utility score (Day 180)**  **N=71**  **β (95% CI)**  **p value** | **VAS score (Baseline)**  **N=210**  **β (95% CI)**  **p value** | **VAS score (Baseline +UVA)**  **N=190**  **β (95% CI)**  **p value** | **VAS score (Day 180)**  **N=27**  **β (95% CI)**  **p value** |
| --- | --- | --- | --- | --- | --- | --- |
| **Age** (years) | -0.002 (-0.006, 0.001); p=0.186 | - 0.002 (-0.006, 0.002); p=0.379 | -0.006 ( -0.010, 0.001); p= 0.013 | -0.257 ( -0.519, 0.006); p= 0.055 | -0.357 ( -0.648, -0.067); p= 0.016 | -0. 118 ( -1.196, 0.961); p= 0.819 |
| **Sex** (ref female) |  |  |  |  |  |  |
| Male | 0.089 (0.000, 0.177); p=0.050 | 0.088 (-0.007, 0.183); p=0.069 | 0.014 ( -0.098, 0.126); p= 0.799 | 6.564 (0.264, 12.865); p= 0.041 | 7.767 (1.092, 14.442); p= 0.023 | 1.130 ( -15.820, 18.080); p= 0.888 |
| **HIV status** (ref negative) |  |  |  |  |  |  |
| Positive | -0.080 (-0.168, 0.007); p=0.073 | - | -0.035 ( -0.150, 0.080); p= 0.546 | 1.345 ( -4.966, 7.655); p= 0.675 | - | -6.563 ( -22.423, 9.296); p= 0.390 |
| Unknown | 0.138 (-0.047, 0.323); p=0.144 | - | 0.042 ( -0.174, 0.257); p= 0.700 | 11.663 ( -2.194, 25.520); p= 0.099 | - | -9.529 ( -50.665, 31.607); p= 0.627 |
| **Active TB status** (Ref no) |  |  |  |  |  |  |
| Yes | -0.084 (-0.191, 0.023); p=0.123 | -0.104 ( -0.214, 0.006); p=0.063 | -0.013 ( -0.121, 0.096); p= 0.818 | -4.621 ( -12.286, 3.043); p= 0.236 | -3.386 ( -11.133, 4.362); p= 0.390 | -19.897 ( -40.826, 1.031); p= 0.061 |
| **Education status** (Ref college or higher |  |  |  |  |  |  |
| No formal schooling | -0.008 (-0.226, 0.210); p=0.943 | -0.063 ( -0.306, 0.180); p= 0.608 | 0.190 ( -0.029, 0.409); p= 0.088 | -7.033 ( -22.213, 8.147); p= 0.362 | -10.806 ( -27.293, 5.681); p= 0.198 | - |
| Primary incomplete or complete | 0.041 (-0.109, 0.190); p= 0.592 | 0.027 ( -0.135, 0.189); p= 0.744 | 0.033 ( -0.141, 0.207); p= 0.705 | -2.917 ( -13.763, 7.930); p= 0.596 | -3.486 ( -15.024, 8.052); p= 0.552 | 0.094 ( -31.088, 31.277); p= 0.995 |
| Secondary school complete | 0.047 (-0.110, 0.204); p= 0.553 | 0.026 ( -0.144, 0.196); p= 0.763 | 0.064 ( -0.126, 0.254); p= 0.501 | 1.931 ( -9.499, 13.360); p= 0.739 | 0.791 ( -11.369, 12.951); p= 0.898 | -3.090 ( -42.525, 36.345); p= 0.869 |
| Some secondary education | -0.014 (-0.170, 0.141); p= 0.858 | -0.038 ( -0.209, 0.134); p=0.666 | 0.148 ( -0.049, 0.345); p= 0.138 | -0.790 ( -12.008, 10.427); p= 0.890 | -2.966 ( -15.057, 9.125); p= 0.629 | 7.756 ( -23.058, 38.570); p= 0.598 |
| **Occupation** (Ref currently employed) |  |  |  |  |  |  |
| Retired | 0.437 ( -0.092, 0.966); p= 0.105 | 0.495 (-0.052, 1.043); p= 0.076 | - | -19.155 ( -59.026, 20.717); p= 0.345 | -16.578 ( -57.270, 24.115); p= 0.422 | - |
| Self-employed | -0.062 (-0.175, 0.051); p= 0.278 | -0.034 ( -0.157, 0.088); p= 0.580 | 0.037 ( -0.096, 0.170); p= 0.581 | -2.034 ( -9.925, 5.857); = 0.612 | -0.622 ( -8.966, 7.723); p= 0.883 | 2.841 ( -21.942, 27.624); p= 0.809 |
| Student | -0. 044 (-0.200, 0.113); p= 0.584 | -0.001 ( -0.173, 0.172); p= 0.995 | 0.006 ( -0.205, 0.217); p= 0.957 | 1.489 ( -9.497, 12.476); p= 0.789 | 0.704 ( -11.074, 12.481); p= 0.906 | -15.999 ( -53.609, 21.610); p= 0.377 |
| Unemployed | 0.034 (-0.080, 0.149); p= 0.556 | 0.046 ( -0.077, 0.168); p= 0.461 | 0.008 ( -0.132, 0.148); p= 0.908 | 3.284 ( -4.863, 11.431); p= 0.428 | 3.447 ( -5.062, 11.956); p= 0.425 | -6.633 ( -30.431, 17.166); p= 0.560 |
| **Days unwell before admission** | -0.001 (-0.002, -0.000); p= 0.013 | -0.002 ( -0.003 -0.000); p= 0.039 | -0.001 ( -0.001, -0.000); p= 0.003 | -0.040 ( -0.089, 0.010); p= 0.114 | -0.033 ( -0.141, 0.076); p= 0.553 | -0.057 ( -0.603, 0.490); p= 0.827 |
| **UVA** | - | -0.025 ( -0.051 - 0.002) P= 0.068 | - | - | -0.569 ( -2.434 - 1.295)P= 0.547 | - |

S2 Figure 1: Mean EQ-5D utility scores across groups at different time points

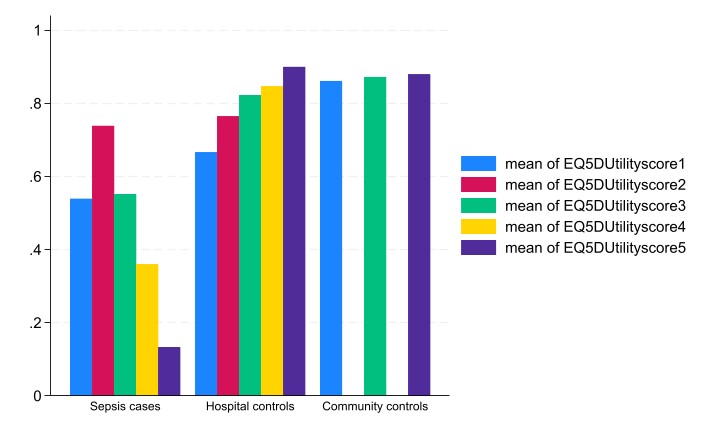

Note: EQ5DUtilityscore1 = Mean utility score at Baseline (Day 0), EQ5DUtilityscore2 = Mean utility score at Day 7, EQ5DUtilityscore3 = Mean utility score at Day 28, EQ5DUtilityscore4 = Mean utility score at Day 90 and EQ5DUtilityscore5 = Mean utility score at Day 180. This figure presents the mean EQ-5D utility scores at five time points (Baseline, Day 7, Day 28, Day 90, and Day 180) for sepsis cases, hospital controls, and community controls

S2 Figure 2: Mean EQ-5D VAS scores across groups at different time points

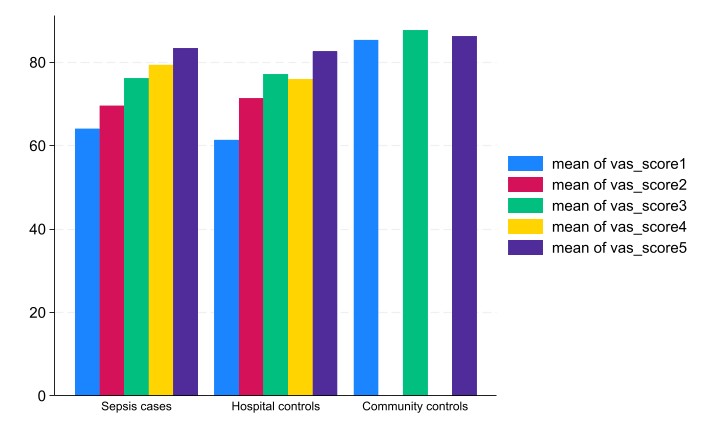

Note: vas_score1 = Mean VAS score at Baseline (Day 0), vas_score2 = Mean VAS score at Day 7, vas_score3 = Mean VAS score at Day 28, vas_score4 = Mean VAS score at Day 90 and vas_score5 = Mean VAS score at Day 180. This figure presents the mean EQ-5D VAS scores at five time points (Baseline, Day 7, Day 28, Day 90, and Day 180) for sepsis cases, hospital controls, and community controls.
