## Supplementary material for "Health-related quality of life among patients with sepsis in Blantyre, Malawi: an observational cohort study": Study Checklist

**S1 Checklist for planning studies including HRQoL measures CHEST 2020; 158(1S): S49-S56**

| No | Item | Response | Location |
| --- | --- | --- | --- |
| 1 | Develop a sound rationale for inclusion of HRQOL | We have indicated a sound rationale | Introduction |
| 2 | Develop hypotheses for specific HRQOL domain(s) | We developed a hypothesis for HRQoL associated with sepsis | Introduction |
| 3 | Select a validated instrument or develop and assess the psychometric properties of a new instrument | We selected validated tool and used by other studies including a SSA context and suited for the clinical condition under study (Sepsis a syndromic condition with multiple underlying aetiologies) where a generic measure was appropriate than disease specific | Introduction and methods |
| 4 | Understand the relevant HRQOL items and domains | Thorough reading from EuroQol resources. Summary presented in the manuscript | Methods |
| 5 | Plan the HRQOL survey administration and timing | Survey administration plan was done and explained in this manuscript | Methods - study recruitment procedures |
| 6 | Track compliance and missing data | Compliance and missing data were tracked | Methods |
| 7 | Plan for the scoring and interpretation of the instrument | Plan was made and summarised | Methods - data handling and statistical analysis |
| 8 | Develop a statistical analysis plan, including how missing data will be handled and subgroup or sensitivity analyses will be performed | Statistical analysis was planned to include handling of missing data and sensitivity analysis conducted | Methods - data handling, statistical analysis and reported in supplementary material |
| 9 | Plan how to clearly present HRQOL data | Plan was in place for presenting data | Methods – statistical analysis |
| 10 | Determine the clinical significance and generalizability of the findings | Clinical significance determined and generalisability reported | Discussion and conclusion |
| 11 | Consider potential limitations, including potential biases from proxy respondents and missing data | Limitations identified and reported | Discussion -Limitations of the study |
